# Immunogenomic intertumor heterogeneity across primary and metastatic sites in a patient with lung adenocarcinoma

**DOI:** 10.1101/2021.08.01.21260883

**Authors:** Runzhe Chen, Jun Li, Junya Fujimoto, Xin Hu, Kelly Quek, Ming Tang, Akash Mitra, Carmen Behrens, Chi-Wan Chow, Peixin Jiang, Latasha D. Little, Curtis Gumbs, Xingzhi Song, Jianhua Zhang, Dongfeng Tan, John V. Heymach, Ignacio Wistuba, P. Andrew Futreal, Don L. Gibbons, Lauren A. Byers, Jianjun Zhang, Alexandre Reuben

**Affiliations:** Department of Thoracic/Head and Neck Medical Oncology, the University of Texas MD Anderson Cancer Center, Houston, Texas 77030, USA; Department of Genomic Medicine, the University of Texas MD Anderson Cancer Center, Houston, Texas 77030, USA; Department of Translational Molecular Pathology, the University of Texas MD Anderson Cancer Center, Houston, Texas 77030, USA; Department of Pathology, the University of Texas MD Anderson Cancer Center, Houston, Texas 77030, USA

**Author notes:** co-first authors (Runzhe Chen, Jun Li). Correspondence (D.L.G.), (L.A.B.), (J.J.Z.), (A.R.).

**Keywords:** lung adenocarcinoma, intertumor heterogeneity, genomic, T cell repertoire

## Abstract

**Background:** Lung cancer is the leading cause of cancer death, partially owing to its extensive heterogeneity. The analysis of intertumor heterogeneity has been limited by an inability to concurrently obtain tissue from synchronous metastases unaltered by multiple prior lines of therapy.

**Methods:** In order to study the relationship between genomic, epigenomic and T cell repertoire heterogeneity in a rare autopsy case from a young female never-smoker with late-stage lung adenocarcinoma (LUAD), we did whole-exome sequencing (WES), DNA methylation and T-cell receptor (TCR) sequencing to characterize the immunogenomic landscape of one primary and 19 synchronous metastatic tumors.

**Results:** We observed heterogeneous mutation, methylation, and T cell patterns across distinct metastases including a set of prevalent T cell clonotypes which were completely excluded from left-side thoracic tumors. Though a limited number of predicted neoantigens were shared, these were associated with homology of the T cell repertoire across metastases. Lastly, ratio of methylated neoantigen coding mutations was negatively associated with T-cell density, richness and clonality, suggesting neoantigen methylation may partially drive immunosuppression.

**Conclusions:** Our study demonstrates heterogeneous genomic and T cell profiles across synchronous metastases and how restriction of unique T cell clonotypes within an individual may differentially shape the genomic and epigenomic landscapes of synchronous lung metastases.

## Background

Lung cancer is the leading cause of cancer death, partially owing to its extensive heterogeneity [1, 2]. It has been proposed that this extensive heterogeneity results from successive clonal expansion and selection of the fittest clones influenced by genomic accumulation and somatic epigenetic alterations [3–6]. However, tumor evolution may also be shaped by pressure from the immune system, which can prune the most immunogenic branches of the tumor [7].

T cells play a crucial role in preventing cancer development through antigen-specific detection and destruction of malignant cells, though evolving tumors can eventually escape immune surveillance through a process termed immunoediting [4, 8–11]. Few studies have addressed the impact of the T cell repertoire in shaping metastatic heterogeneity [12–14] with most work to date evaluating longitudinal changes spanning multiple timepoints and therapies. Though these studies offer crucial insights, they do not allow the evaluation of intrinsic intertumor heterogeneity in absence of selective pressure from therapy. Furthermore, to date, the analysis of intertumor heterogeneity has been limited by an inability to concurrently obtain tissue from synchronous metastases unaltered by multiple prior lines of therapy [15–17].

Here, we sought to study the relationship between genomic, epigenomic and T cell repertoire heterogeneity in a rare autopsy case from a female never-smoker in her early 30s with late-stage lung adenocarcinoma (LUAD) with more than 20 synchronous metastases. We observed heterogeneous mutation, methylation, and T cell patterns across distinct metastases including a set of prevalent T cell clonotypes which were completely excluded from left-side thoracic tumors. Our work further highlights neoantigen methylation as a potential mechanism driving immunosuppression and some of the hurdles facing the treatment of late-stage lung cancer.

## Methods

### Human subject research

We collected 20 tumor samples and one normal GI sample at autopsy. Collection and use of patient samples were approved by the Institutional Review Board of the University of Texas MD Anderson Cancer Center. Clinical information is presented in **Supplementary Table 1**.

### Sample collection

DNA of collected samples was isolated from FFPE tissues using the AllPrep DNA/RNA FFPE Kit (Qiagen, Hilden, Germany). Hematoxylin and eosin (H&E) slides of each case were reviewed by experienced lung cancer pathologists under the microscope to assess the percentage of pre-/micro-invasive neoplastic lesions and tumor tissues versus normal tissues. Tumor cell viability was also assessed by examining the presence of necrosis in the tissues. Only samples with enough viable tumor cells were selected for WES, methylation and immunoSEQ.

### DNA preparation

Unstained tissue sections (10μm thick) were deparaffinized in xylene and 100% ethanol (twice in each for 10 minutes). The macrodissected tumor areas of the deparaffinized tissues were placed into a 1.5 mL collection tubes for DNA extraction. The tissue was next suspended with Buffer PKD and Proteinase K from the Allprep FFPE kit. After incubating at 56 °C for 15 min then on ice for 5 min, the mixed solution was centrifuged for 15 minutes at 20,000 x g. Finally, the DNA samples were quantified by Nano Drop 1000 Spectrophotometer (Thermo Scientific, Wilmington, DE, USA). The fragmentation sizes were evaluated by the Agilent 2200 Tape Station system using the Genomic DNA Screen Tape Assay (Agilent Technologies, Santa Clara, CA, USA).

### Whole-exome sequencing

Exome capture was performed on 500ng of genomic DNA per sample based on KAPA library prep (Kapa Biosystems) using the Agilent SureSelect Human All Exon V4 kit according to the manufacturer’s instructions and paired-end multiplex sequencing was performed on the Illumina HiSeq 2000 sequencing platform. The average sequencing depth was 178x (ranging from 63x to 225x, standard deviation +/- 31).

### Mutation calling

Tumor contents and major/minor copy number changes were estimated by Sequenza (v2.1.2) [18]. To control those FFPE caused artifact contaminations, somatic single nucleotide variants (SNVs) was first called using MuTect version 1.1.4 [19], VarScan 2 [20] and Strelka2 [21] with default setting, respectively. Then, the following filtering criteria were applied to each callers: 1) sequencing depth ≥20× in tumor DNA and ≥10× in germline DNA; and 2) variant allele frequency (VAF) ≥ 0.02 in tumor DNA and < 0.01 in germline DNA; and 3) the total number of reads supporting the variant calls is ≥4; and 4) variant frequency is < 0.01 in ESP6500, 1000 genome and EXAC databases; Those mutations called by Mutect with a LOD score<10 was further filtered out. Those mutations called by Strelka with a quality score below 35 was also filtered out. Finally, only those mutations were kept if called by any of the two tools and rescued if any were rejected but shared by at least two tumors. Identified missense mutations were manually reviewed using the Integrative Genomics Viewer version 2.3.61 [22, 23].

### Phylogenetic analysis

Ancestors were germ line DNA assuming with no mutations. The phylogenetic tree was generated as described [24]. A binary presence/absence matrix of all somatic mutations detected was used as input for R package phangorn version 2.0.2 [25].

### Neoantigen predictions

Nonsynonymous mutations were identified from WES profiling and the binding affinity with patient-restricted MHC Class I molecules of all possible 9- and 10-mer peptides spanning the nonsynonymous mutations was evaluated with the NetMHC3.4 algorithm based on HLA-A, HLA-B, and HLA-C alleles of each patient [26–28]. Candidate peptides were considered as HLA binders when IC50<500 nM with high affinity binders presenting IC50<50 nM.

### DNA methylation profiling and tumor–immune microenvironment deconvolution

Genomic DNA (approximately 500 ng) was bisulfite converted using EZ DNA Methylation Kit (Zymo Research Corp. Irvine, CA, USA) following the manufacturer’s protocol. Bisulfite converted DNA materials were then processed and hybridized to the Infinium Human Methylation 450k arrays (Illumina, San Diego, CA, USA) according to manufacturer’s recommendation. Preprocessing and initial quality assessments of the raw data were examined using the following R packages. Subset-quantile within-array normalization (SWAN) [29] was used to normalize raw methylation values. IlluminaHumanMethylation450k.db annotation package was used to annotate the CpG probes location. Before any genomics and statistical analyses were conducted, we normalized and inspected the methylation data for the presence of substantial confounding batch effects. Cellular deconvolution analyses were carried out using estimated cellular fractions using MethylCIBERSORT [30].

### TCRβ sequencing and comparison parameters

Immunosequencing of the CDR3 regions of human TCRβ chains was performed using the protocol of immunoSEQ (Adaptive Biotechnologies, hsTCRβ Kit) [31–33]. Two sets of PCRs were performed on DNA extracted from the tissues collected. The initial PCR used a mix of multiplexed V- and J-gene primers which amplify all possible recombined receptor sequences from the DNA sample, and then a second PCR designing to add unique DNA barcodes to each PCR product was followed. After that, samples were pooled together with a negative and a positive control. The pools were then sequenced on an Illumina MiSeq platform using 150 cycle paired-end protocol and sequence-ready primers. After finish the sequencing, the raw data were transferred to Adaptive Biotechnologies and processed into a report including those passed quality-check samples and a normalized and annotated TCRβ profile repertoire accordingly. Profile of TCR rearrangements is presented in **Supplementary Data**.

T-cell density in FFPE tissue samples was calculated by normalizing TCR-β template counts to the total amount of DNA usable for TCR sequencing, where the amount of usable DNA was determined by PCR-amplification and sequencing of housekeeping genes expected to be present in all nucleated cells. T-cell richness is a metric of T cell diversity, and it is calculated by on the T-cell unique rearrangements. T-cell clonality is a metric of T cell proliferation and reactivity, and it is defined as 1-Peilou’s evenness. Clonality ranges from 0 to 1: values approaching 0 indicate a very even distribution of frequency of different clones (polyclonal), whereas values approaching 1 indicate a distinct asymmetric distribution in which a few activated clones are present at high frequencies (monoclonal). Statistical analysis was performed in R version 3.2. JI is conceptually a percentage of how many objects of two sets have in common out of how many objects they have in total. JI = (number of rearrangements in common) / (total number of rearrangements)

### Statistical Analysis

Graphs were generated with GraphPad Prism 8.0. Spearman’s rank correlations were calculated to assess the association between 2 continuous variables. Kruskal-Wallis tests were used for categorical variables with more than 2 levels. P-values less than 0.05 were considered to be statistically significant.

## Results

### Patient information

A female never-smoker in her early 30s presented to her primary care physician complaining of weakness in her upper right arm lasting for two weeks. Physical examination was unremarkable, other than grade 3 weakness in her right upper limb. Shortly after, she was hospitalized due to acute venous thromboembolism of this arm. She was started on anti-coagulants and underwent computed tomography (CT) scans of the chest, abdomen and pelvis and magnetic resonance imaging (MRI) of the brain as part of the work up. Numerous nodules were detected suggestive of extensive metastasis and a 1.9 cm left lung mass was consistent with a lung primary (**Fig. 1a-b**). A liver biopsy revealed the diagnosis of poorly differentiated LUAD. The patient underwent palliative radiation therapy for C5-C7 spine metastases with 10×30 cGy and one dose of chemotherapy with carboplatin and paclitaxel while awaiting molecular profiling results. Her condition deteriorated rapidly and she expired 13 days following her sole dose of chemotherapy. An autopsy was performed and widely disseminated metastatic carcinoma involving multiple systems and organs was observed.

**Figure 1.**
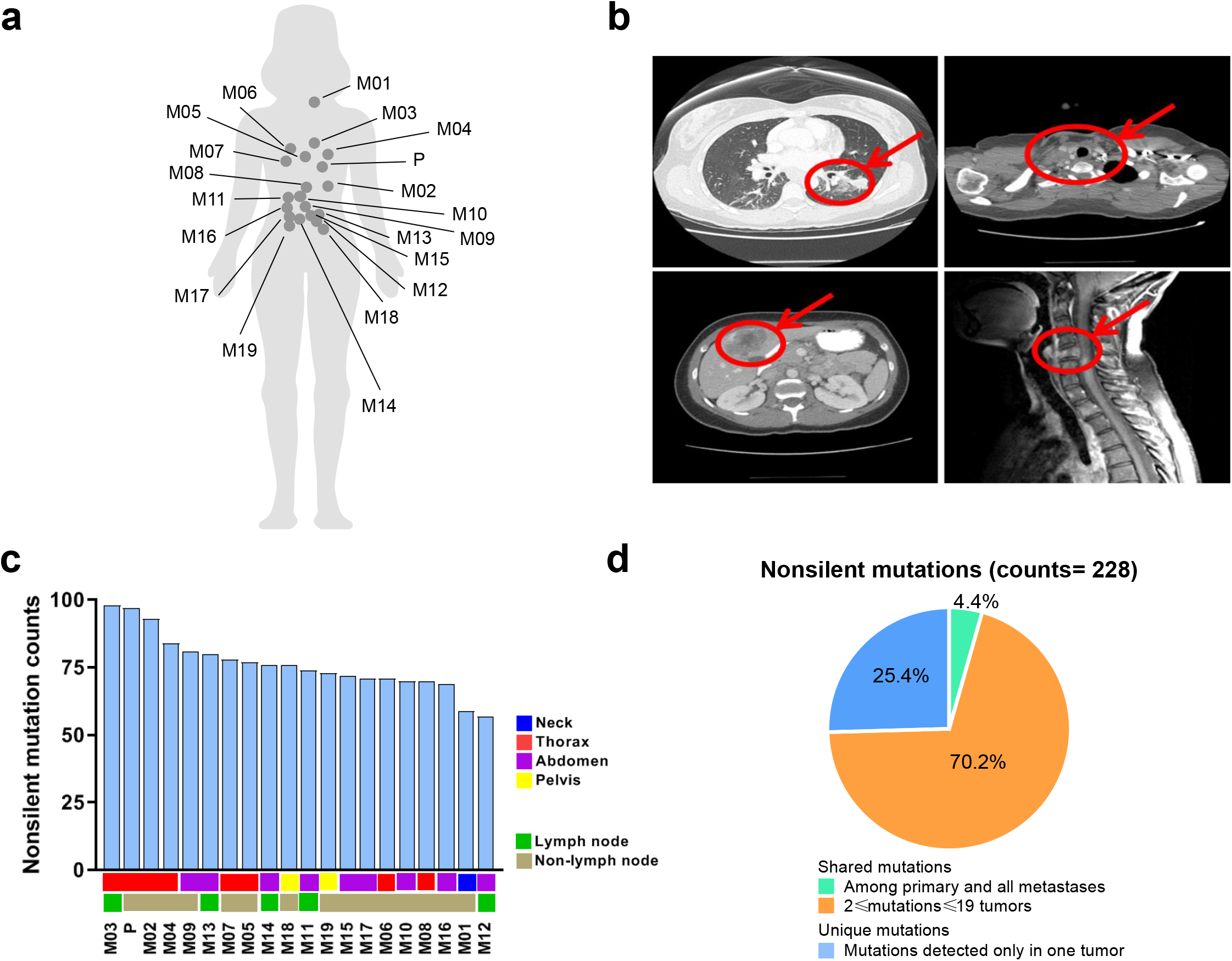
Synchronous metastatic tumors exhibit heterogeneous growth and somatic mutation and neoantigen patterns. ) Anatomical map of representative biospecimen collection sites in the patient. **b)** Representative CT and MRI scans of different resected tumors: primary lung tumor, cervical lymph node enlargement, liver metastasis and C5 spine compression fracture by metastasis. **c)** Non-silent mutation counts in tumors. **d)** Fraction of shared and unique non-silent mutations across tumors.

To understand the genomic and T cell landscape of this extensively metastatic LUAD, 20 tumor samples (**Supplementary Table 1**) including the left lung (primary tumor, P), thyroid gland (M01), left pleural cavity (M02), left hilar lymph node (M03), left-side parietal pleura (M04), heart (M05), right lung (M06), right pleural cavity (M07), 12^th^ thoracic vertebra (M08), gastrointestinal (GI) tract (M09), liver (M10), 4 abdominal lymph nodes (M11-14), left adrenal gland (M15), two metastases in the right kidney (M16 and M17), left and right ovaries (M18 and M19) as well as one histologically normal sample from the GI tract were collected and subjected to whole-exome sequencing (WES), DNA methylation array and T cell receptor (TCR) sequencing.

### Distinct mutational profiles are seen across primary tumor and synchronous metastases

Overall, 228 non-silent mutations were detected with an average of 76 per sample (range=57-98). The number of non-silent mutations varied between tumors, with only 10 shared across all 20 samples (**Fig. 1c**). Of these non-silent mutations, 170 (75%) were shared by at least two tumors while 58 (25%) were unique (**Fig. 1d**). When canonical cancer gene mutations were analyzed[34–36], commonly-mutated cancer genes included TP53, CDKN2A, ASXL1 and MET in this patient (**Supplementary Fig. 1a**). Only TP53 mutation (chromosome 17_7578382, stop gain, spectrum G->C) was detected in all tumors suggesting TP53 mutation was an early genomic event, while other cancer gene mutations were later events which may have followed subclonal diversification. We also constructed a phylogenetic tree to depict the genomic heterogeneity and evolutionary trajectory of these metastatic tumors. As shown in **Supplementary Fig. 1b**, the phylogenetic structure varied considerably between tumors highlighting profound genomic heterogeneity within this patient. We also utilized the Jaccard index (JI), which takes into consideration the proportion of shared non-silent mutations between any two samples. The JI ranged from 0.14 to 0.82 (average=0.49) with more proximal tumors generally more genetically similar (**Supplementary Fig. 1c**). Homology between the primary and metastases ranged from 0.14 to 0.73 (average=0.33), with the thoracic lesions including the left pleural metastasis, left hilar lymph node and right pleural metastasis exhibiting the greatest similarity with the primary tumor. Taken together, these results reveal marked genomic heterogeneity across different metastases within the same patient.

### The T cell infiltrate in distant metastases is more dense, diverse and reactive

The crucial role of T cells in immunoediting led us to study the T cell repertoire to further investigate the spatial heterogeneity of T-cell responses[37, 38]. T-cell density, an estimate of the fraction of T cells within a tumor, ranged from 3% to 38% (average=13%, **Fig. 2a**), while richness, a measure of T-cell diversity, ranged from 4,168 to 23,487 unique T-cell rearrangements (average=14,344 unique rearrangements, **Fig. 2b**). T-cell clonality, a measure of T-cell reactivity, ranged from 0.02 to 0.05 (average=0.04, **Fig. 2c**). All TCR metrics were positively inter-correlated (Density *vs*. Richness: r=0.53, p=0.02; Density *vs*. Clonality: r=0.54, p=0.02; Clonality *vs*. Richness: r=0.54, p=0.01, **Fig. 2d**). Compared to prior work from our group in an early-stage LUAD cohort[39], lower T-cell density (p<0.05) and clonality (p<0.0001) and higher richness (p<0.0001) were observed in tumors from this patient (**Supplementary Fig. 2a-e**).

**Figure 2.**
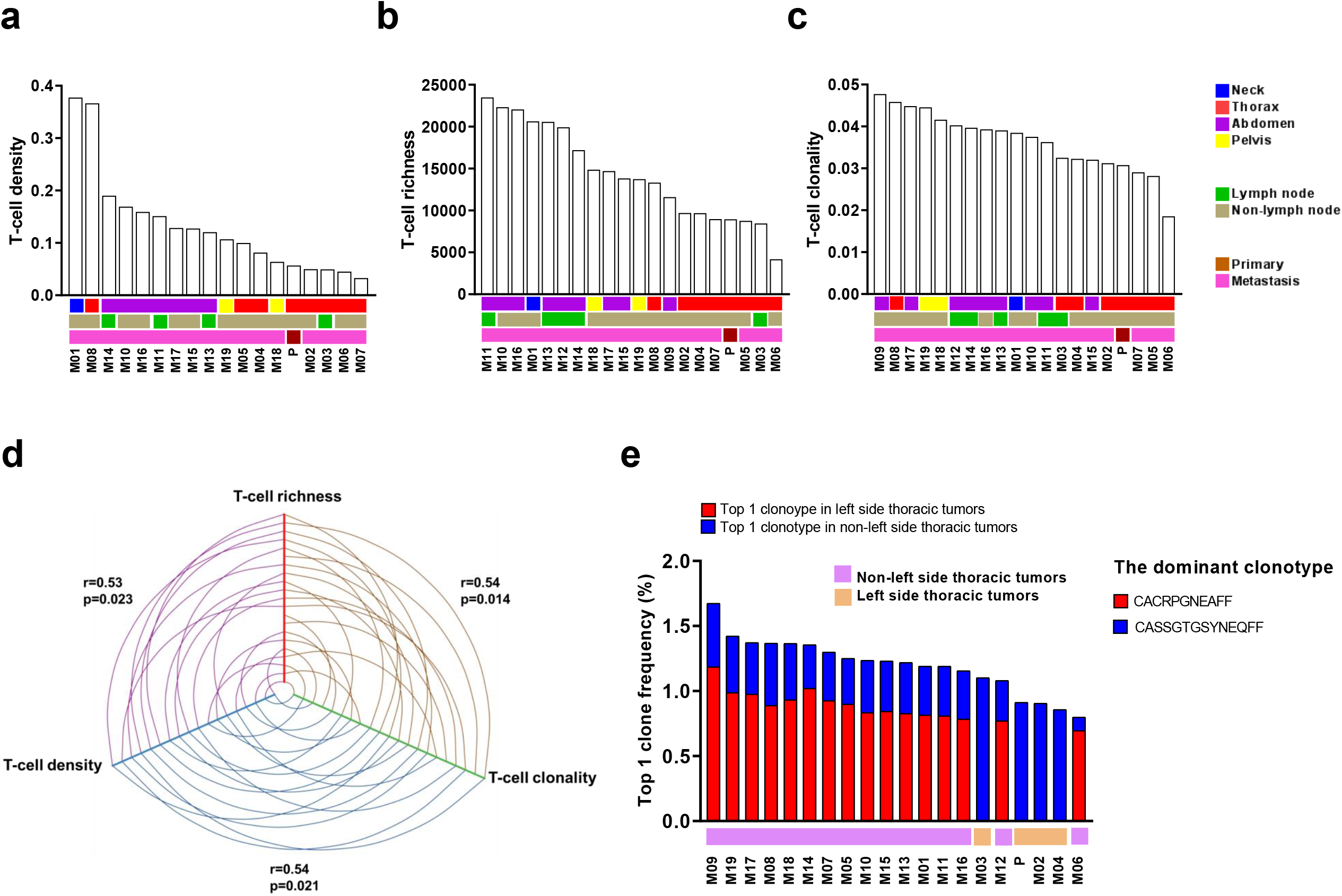
Characterization of TCR repertoire metrics across tumors. T-cell **a)** density, **b)** richness and **c)** clonality. **d)** Correlation between T-cell fraction, richness and clonality. **e)** Distribution of most prevalent TCR clonotype.

Tumors were then grouped anatomically. Non-thoracic tumors displayed higher T-cell density (p<0.01), richness (p<0.0001) and clonality (p<0.01) than thoracic tumors (**Supplementary Fig. 3a-c**), perhaps owing to their anatomical location far from the primary tumor which are more likely to escape from immune surveillance[40, 41]. Lymph nodes serve as sites of T cell priming, activation, and modulation, leading us to speculate that the interaction between metastatic cancer cells and T cells in lymph nodes may be distinct compared to other sites of metastases. However, no statistical differences were observed in relation to lymph node involvement (**Supplementary Fig. 3d-f**). Taken together, these data suggest differences in T cell response based on anatomical site, that is, T cell exclusion, suppression and a more focused T cell response in proximity to the primary tumor.

### Distinct TCR repertoire profiles are associated with left-side thoracic tumors

To evaluate T-cell responses in the tumors, we next focused on the most prevalent TCR clonotypes. Distinct clonotypes were detected in left-side thoracic tumors (left lung tumor, left pleural cavity and rib, left pulmonary hilar lymph node and left side parietal pleura) compared to others. Strikingly, the most prevalent clonotype in “other” tumors (*CACRPGNEAFF*) was entirely undetectable in left-side thoracic tumors (P, M02, M03 and M04) (**Fig. 2e**). Similar trends were also observed among the top 5 and 10 TCR clonotypes with certain clonotypes completely excluded from left side thoracic tumors (**Supplementary Fig. 4a-b**). These data illustrate spatial restriction even among the most prevalent T cell clonotypes across synchronous metastases.

### T-cell repertoire heterogeneity is observed across differentially growing tumors

To gain deeper insights into TCR heterogeneity, we assessed the overlap between T cell repertoires across different tumors. We first compared the proportion (JI) and frequency of T cell clonotypes shared between the primary tumor and metastases. In accordance with the unique T cell clonotype pattern observed in tumors from the left side thorax, proportions and frequencies of shared T cells were much more similar between the three left thoracic metastases (M02, M03 and M04) and primary tumor (P) (**Fig. 3a-c**). T cell repertoire heterogeneity was evident across all tumors, with an average JI value of 0.35 (ranging from 0.12 to 0.61) and more shared T cells between proximal tumors (**Fig. 4a**), significantly higher than in a previously published cohort of 11 multi-region localized non-small cell lung cancers[42] (average 0.35 *vs*. 0.17, p<0.0001) (**Supplementary Fig. 5**). Next, we studied the proportion and frequency of shared T cell clonotypes across all 20 tumors. In total, 599 shared T-cell clones were shared across all tumors, with proportions ranging from 3.0% to 15.4% (average=5.39%) and frequencies accounting for 11.9% to 21.5% of the T cell repertoire (average=15.96%) (**Supplementary Fig. 6a-b**). Of interest, both a greater proportion (p<0.01) and percentage (p<0.01) of shared T cell clones were observed in thoracic tumors compared to non-thoracic tumors (**Supplementary Fig. 6c-d**).

**Figure 3.**
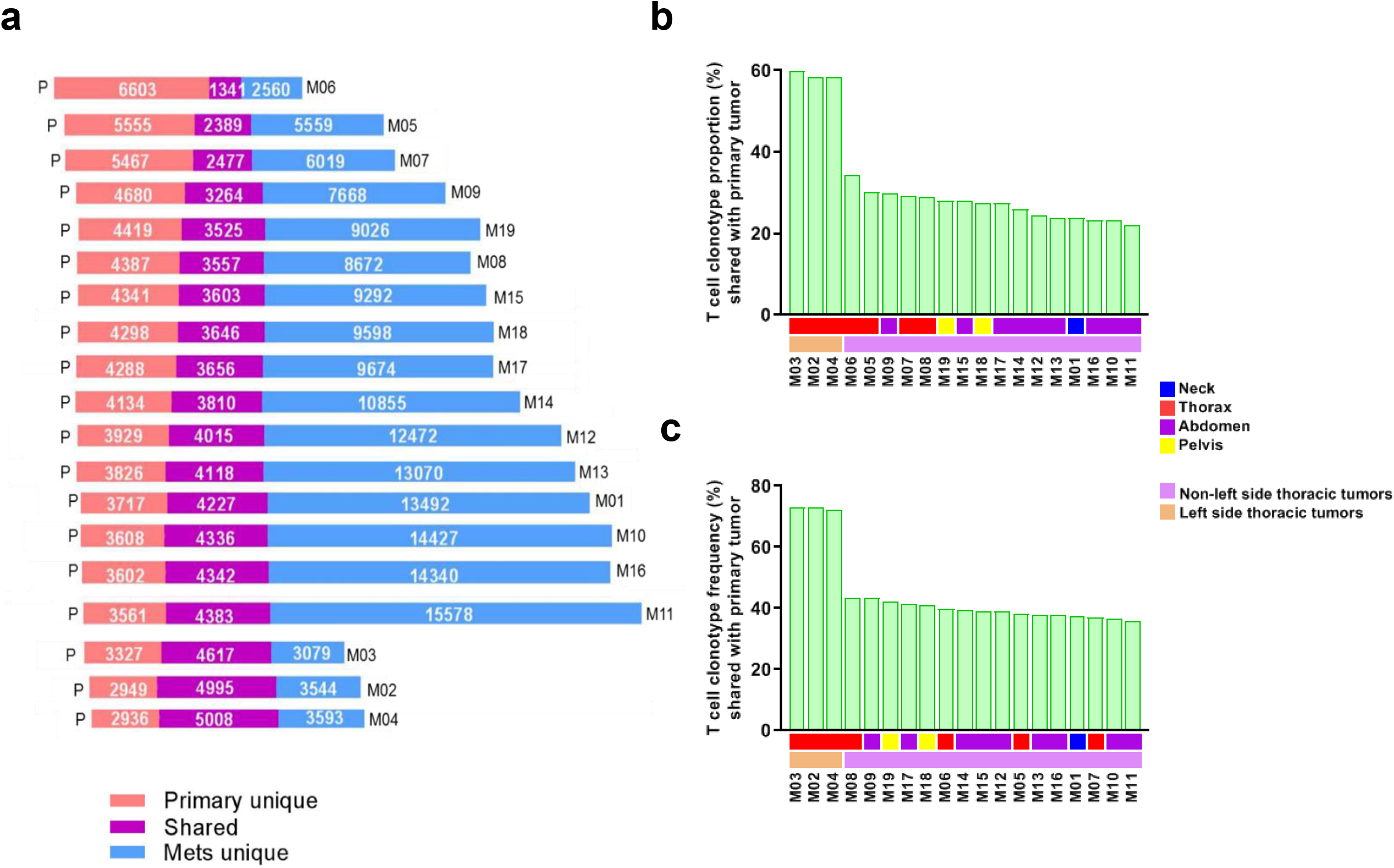
T-cell repertoire heterogeneity is observed across differentially growing tumors. ) Number of T-cell clonotypes in the primary tumor (red), metastases (blue) or shared (purple). **b)** Shared T-cell clonotype proportions and **c)** frequencies between the primary tumor and metastases.

**Figure 4.**
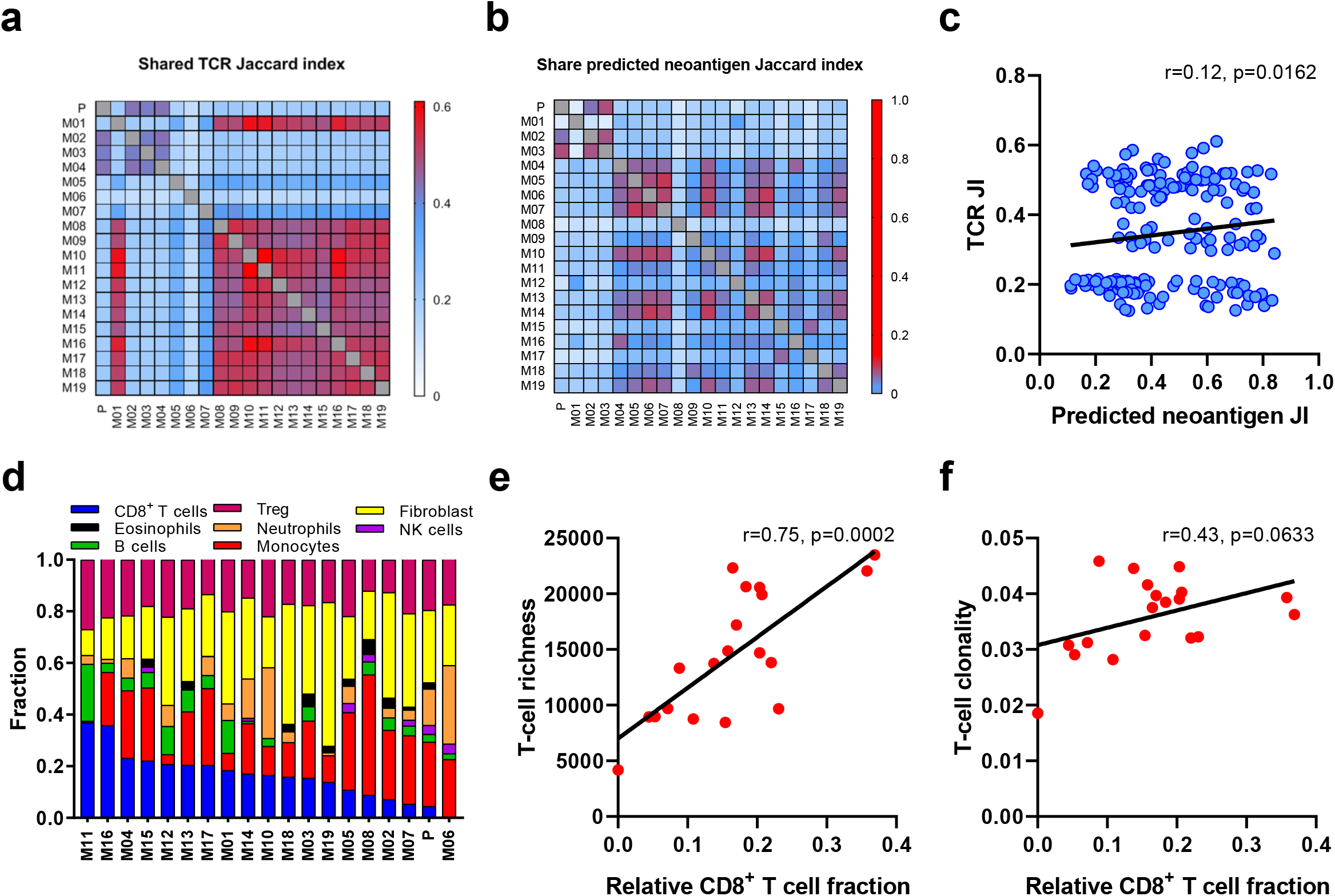
Evolution of synchronous metastases may be shaped by the T cell repertoire. ) Quantification of TCR repertoire heterogeneity across tumors by Jaccard index (JI). **b)** Quantification of predicted neoantigen heterogeneity across tumors by JI. **c)** Correlation between TCR repertoire JI and predicted neoantigen JI. **d)** Deconvolution of immune components and T cell subpopulations by MethylCIBERSORT. **e)** Correlation between T-cell richness and estimated CD8^+^ T-cell fraction. **f)** Correlation between T-cell clonality and estimated CD8^+^ T-cell fraction.

### Evolution of synchronous metastases may be shaped by the T cell repertoire

We next performed *in silico* prediction of HLA-A-, -B-, and -C-presented neoantigens using NetMHC3.4[26–28]. On average, 39 predicted neoantigens (IC50 < 500 nmol/L) were detected per tumor, with the most (n=60) seen in the primary tumor and fewest (n=20) in the thyroid gland. Only 11 high binding affinity neoantigens were detected on average (IC50 < 50 nmol/L) with the most (n=19) also in the primary tumor and least (n=2) in the thyroid gland (**Fig. 5a**). This falls within range but below the average of 53 predicted neoantigens seen in non-smokers from TCGA (**Fig. 5b**). We then evaluated the relationship between the T cell repertoire and predicted neoantigens. Predicted neoantigen heterogeneity was also evident, with the average JI value of 0.44 (ranging from 0.11 to 0.84, **Fig. 4b**), and a weak but statistically-significant positive correlation between T-cell repertoire and neoantigen homology (r=0.12, p=0.0162, **Fig. 4c**), which could suggest the distribution of T cells may be partially driven by their reactivity to shared neoantigens. Interestingly, ratio of methylated neoantigen coding mutations was negatively associated with T-cell density (r=-0.46, p=0.0549), richness (r=-0.55, p=0.0152) and clonality (r=-0.61, p=0.0055) (**Fig. 5c-e**), suggesting neoantigen methylation may contribute to immune suppression and potentially explaining the weak neoantigen associations with T cell repertoire homology.

**Figure 5.**
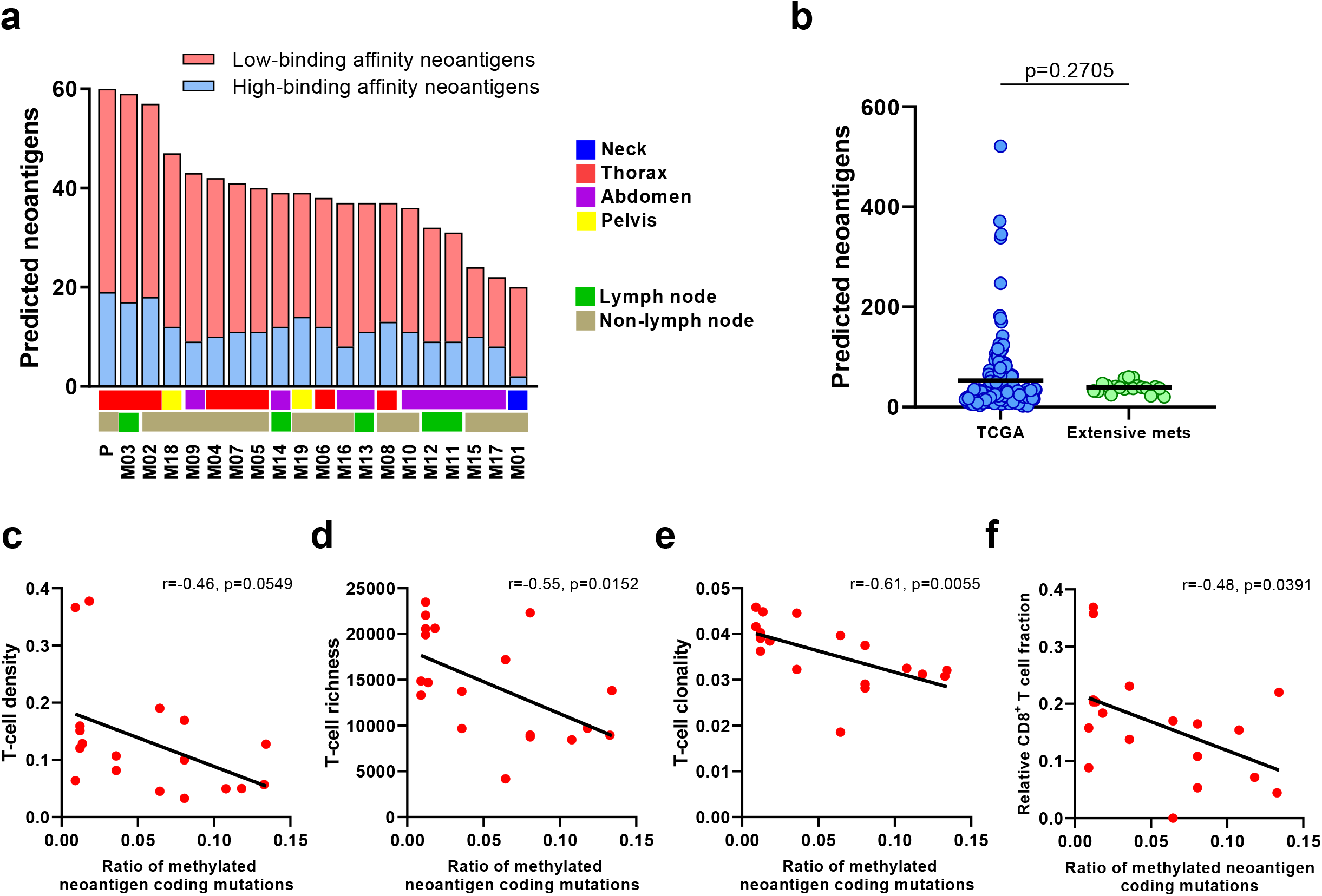
Methylated neoantigen burden is inversely correlated with T cell repertoire metrics. ) Total number of predicted neoantigens across tumors. **b)** Comparison of predicted neoantigens between TCGA cohort and tumors in our study. Correlation between ratio of methylated neoantigen coding mutations and T-cell **c)** density, **d)** richness and **e)** clonality.

In order to classify the T cell subsets infiltrating the tumor, we next performed cellular deconvolution analyses using MethylCIBERSORT (**Fig. 4d**)[30]. Relative CD8^+^ T cell fraction but not CD4^+^ and other immune subsets was proportional to richness (r=0.75, p=0.0002) and clonality (r=0.43, p=0.063) (**Fig. 4e-f**). Furthermore, CD8 to Treg ratio, which correlates with a more favorable outcome in cancer[43, 44], was proportional to T-cell richness (r=0.68, p=0.0012) and clonality (r=0.64, p=0.032) (**Supplementary Fig. 7a-b**). A negative correlation between CD8^+^ T cell fraction and methylated neoantigen coding mutations was also observed (r=-0.48, p=0.0391, **Fig. 5e**). These results highlight the greater proliferative potential of CD8^+^ T cells and suggest T cell reactivity and diversity may be mainly driven by the clonal expansion of CD8^+^ T cells at the patient level, as previously suggested by our group and others in a large cohort of early-stage non-small cell lung cancers[45]. Overall, our findings suggest that the evolution of synchronous metastases may be shaped by the T cell response including in absence of prior therapy.

## Discussion

Metastasis is an evolutionary process shaped by the dynamic interactions between tumor cells and host factors including immune surveillance [46]. T cells play a pivotal role in mediating this process by recognizing antigens presented on MHC molecules at the surface of tumors and carrying out cytotoxic responses against tumor cells harboring their cognate antigens [47]. Accordingly, much importance has been attributed to T cell infiltration in many solid tumors, with more T cells generally associated with a better prognosis [37, 48, 49]. However, recent studies have highlighted the impact of intratumor heterogeneity and bystander T cells [50–52], and suggested that only ∼10% of tumor-infiltrating lymphocytes are capable of recognizing antigens presented by the tumor they have infiltrated [53], prompting deeper investigations into the T cell repertoire. Our understanding of the role of genomic and immune heterogeneity in lung cancer has evolved in recent years, thanks to investigations by our group and others into differences between regions of individual tumors, synchronous metastases and between primary and metastatic tumors [54–58] highlighting potential spatial and temporal factors influencing clinical outcomes [59, 60]. Here, we assess the characteristics of the T cell repertoire in a treatment-naïve non-smoking patient with synchronous lung metastases and depict the interplay between the primary tumor and synchronous metastases [11, 55, 61], revealing extensive immunogenomic intertumor heterogeneity across primary and metastatic sites.

In our study, clonal *TP53* mutations were detectable in all tumors, suggestive of an early genomic event, in line with prior reports [24, 62]. Interestingly, a higher overlap in somatic mutations was observed across proximal tumors suggesting they are more genetically similar, potentially due to metastatic seeding from the primary tumor [63, 64]. Though our study focused on a single patient, the overlap in mutational burden observed between synchronous metastases is in line with previous reports in lung [58], melanoma [65], kidney [66] and colon cancer [67]. Considering the role of somatic mutations in triggering T cell responses through the generation of neoantigens, this overlap suggests these somatic mutations may serve as potential therapeutic targets for vaccination or T cell engineering through targeting of unifying antigens present across all synchronous tumors. This is supported by the modest but significant correlation between shared mutations and shared TCRs though additional studies are needed to confirm these hypotheses.

We observed lower T cell repertoire heterogeneity across synchronous metastases in our study than in our prior work assessing multi-region ITH in localized LUAD [55]. This difference could highlight the distinct resistance mechanisms at play in accelerated progression in our study versus more gradual progression in early-stage LUAD which may have allowed for divergent genomic evolution and immune editing over decades. This is reinforced by the absolute restriction of certain T cell clonotypes to metastases surrounding the primary tumor, which could be reflective of the distinct antigenic environments established in distal tumors. Unfortunately, our lack of deep immune phenotyping data precludes our ability to further investigate the role the distinct immune microenvironments, including chemokine gradients and receptors, may have played in establishing these vastly distinct T cell microenvironments. However, the presence of shared T cell clonotypes could also be indicative of common responses against unifying antigens displayed across synchronous metastases.

Aberrant methylation is involved in tumorigenesis in a variety of cancers [68, 69] and could affect immune surveillance directly by regulating the expression of immune-related genes and/or potential neoantigens[70]. DNA methylation-induced silencing of genes that encode tumor associated antigens form a complex gene-regulatory network for suppressing anti-tumor immune responses and therefore facilitating immune evasion [71]. Interestingly, in the current study, ratio of methylated neoantigen coding mutations was negatively associated with T-cell density, richness and clonality, even at the individual tumor level. One could therefore hypothesize that therapeutic agents modulating methylation could potentially reprogram the immune microenvironment and could exhibit some potential in treating these tumors.

Our study does exhibit certain limitations, including its focus on a single patient. However, analysis of several synchronous tumor sites from a single patient with advanced disease in absence of heavy pre-treatment is rarely possible, mainly due to the lack of clinical indication. Unfortunately, deeper analysis of underlying mechanisms, immune cells and soluble factors influencing T cell trafficking and heterogeneity remain unclear due to the archival nature of these samples and will require further investigation. Despite these limitations, our study provides important evidence of differential tumor-immune responses co-existing in metastases within the same individual, related not only to molecular alterations. As a result, our findings may also partially explain the challenge of treating late-stage lung cancer due to the heterogeneity of metastases. Additional genomic, transcriptomic and immune studies in patients with synchronous metastases could help shed light on these and other mechanisms at play and provide therapeutic insights into late-stage non-small cell lung cancer. Lastly, our findings emphasize the need to obtain multiple biopsies when feasible in patients with multiple synchronous lung tumors as has been suggested in metastatic melanoma, even in absence of selective pressure from therapy. Concurrent analysis of multiple tumors from the same patient could help identify unifying antigens and mechanisms capable of prolonging survival in patients with metastatic lung cancer [65].

## Conclusion

In summary, in this study, we present the immunogenomic landscape of one primary tumor and 19 synchronous metastases from a minimally-pretreated young female never-smoker with late-stage LUAD. Fewer less diverse and reactive T cells infiltrated the metastases nearest to the primary tumor, and a set of prevalent T cell clonotypes were excluded from left-side thoracic tumors further suggesting immune escape near the primary site. Furthermore, shared predicted neoantigens were associated with homology of the T cell repertoire across metastases. Lastly, ratio of methylated neoantigen coding mutations was negatively associated with T-cell density, richness and clonality, suggesting neoantigen methylation may partially drive immunosuppression. Our study demonstrates heterogeneous genomic and T cell profiles across synchronous metastases and how restriction of unique T cell clonotypes within an individual may differentially shape the genomic and epigenomic landscapes of synchronous lung metastases.

## Data Availability

Data are available from the authors upon reasonable request

## Abbreviations

CT: computed tomography
H&E: hematoxylin and eosin
JI: Jaccard index
LUAD: lung adenocarcinoma
MRI: magnetic resonance imaging
SNVs: single nucleotide variants
SWAN: subset-quantile within-array normalization
TCR: T cell receptor
WES: whole-exome sequencing
VAF: variant allele frequency

## Declarations

### Ethics approval

Collection and use of patient samples were approved by the Institutional Review Board of the University of Texas MD Anderson Cancer Center.

### Consent for publication

Not applicable

### Author contributions

A.R., D.L.G., L.A.B. and J.J.Z. conceived the study. J.L. and R.C. led the data analysis. J.F, D.T., and C.W.C. led the pathological assessment, multi-region sample preparation and DNA extraction. R.C., K.Q. and M.T. collected resected specimens and clinical data. L.L. and C.G. performed DNA preparation and whole-exome sequencing. X.S. and J.H.Z. performed sequencing raw data processing. J.L., X.H., K.Q., M.T. and A.M. performed downstream bioinformatics analyses. R.C., C.B, P.J., J.V.H., I.W., P.A.F., D.L.G, L.A.B., J.J.Z. and A.R. interpreted the data for clinical and pathological correlation. R.C. and J.L. performed statistical analyses. R.C., J.J.Z and A.R. wrote the paper. All authors edited the manuscript.

## Acknowledgements

This work was supported by Conquer Cancer Foundation ASCO Young Investigator Award, MD Anderson Physician Scientist Award, Cancer Prevention & Research Institute of Texas (CPRIT) Multiple Investigator Award, TJ Martell Foundation, NIH/NCI R01-CA207295, NIH/NCI U01-CA213273 and Department of Defense (LC170171). A.R. is supported by the Exon 20 Group, Rexanna’s Foundation for Fighting Lung Cancer, the Waun Ki Hong Lung Cancer Research Fund, MD Anderson’s Lung Cancer Moon Shot and the University Cancer Foundation via the Institutional Research Grant program at the University of Texas MD Anderson Cancer Center.

