## Supplementary figures for "Immunogenomic intertumor heterogeneity across primary and metastatic sites in a patient with lung adenocarcinoma"

Chen *et al*.

**Supplementary Figure 1**

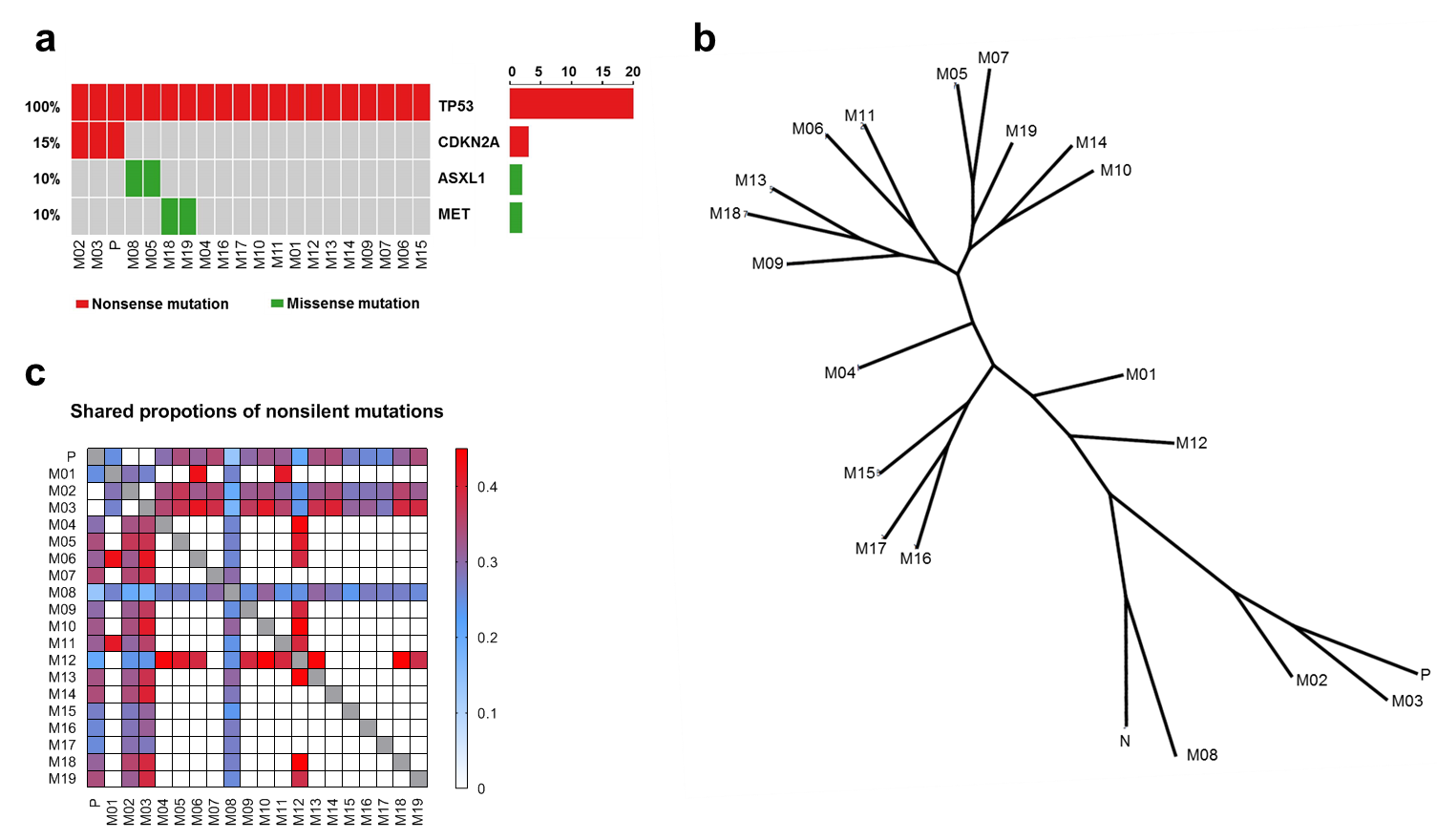

**Supplementary Figure 1. Distinct mutational profiles are seen across primary tumor and synchronous metastases. a)** Oncomap of 20 tumors in the patient with known cancer gene mutations; **b)** Phylogenetic trees were generated from all SNVs by using the Wagner parsimony method in “phangorn” package. Trunk and branch lengths are proportional to the numbers of mutations acquired on the corresponding trunk or branch; **c)** Share proportions of nonsilent mutations quantified by Jaccard index (JI).

**Supplementary Figure 2**

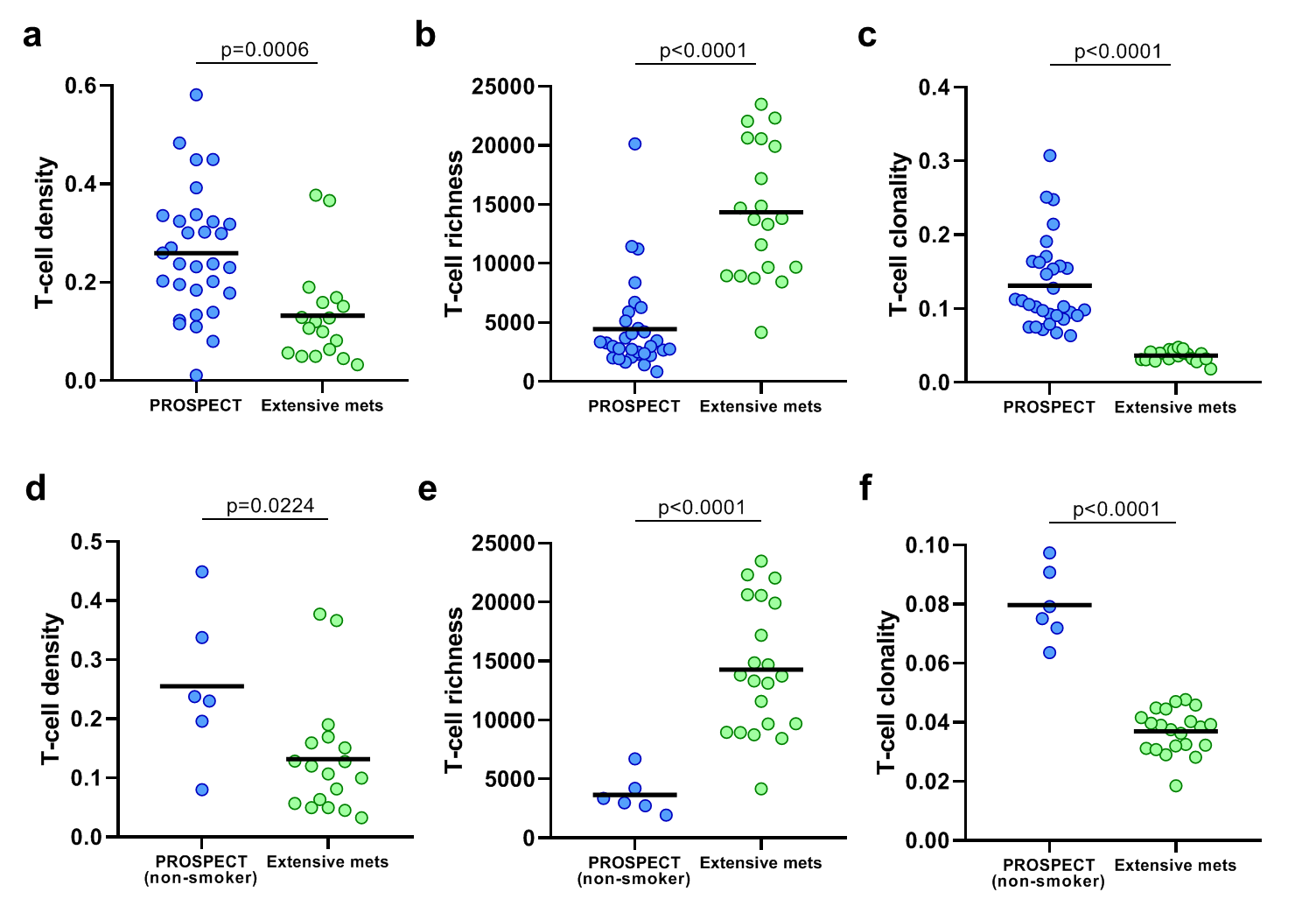

**Supplementary Figure 2. Distinct T cell repertoire metrics between tumors from patients in localized non-small cell lung cancer (NSCLC, PROSPECT cohort) and our patient.** Comparisons of T-cell **a)** density, **b)** richness and **c)** clonality.

**Supplementary Figure 3**

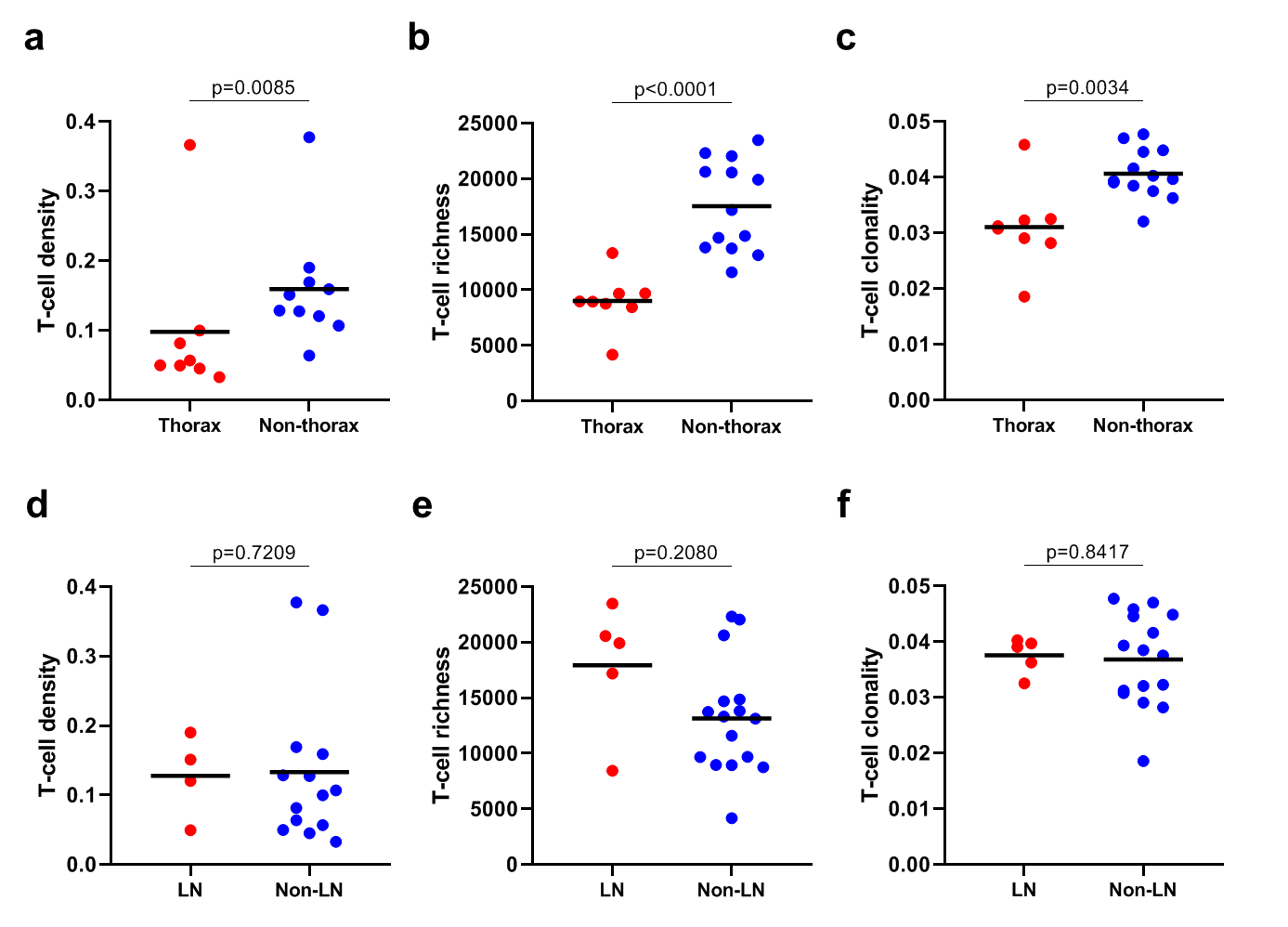

**Supplementary Figu****re 3. Distinct T cell repertoire metrics between thoracic and non-thoracic tumors but no differences in relation to lymph node involvement.** T-cell **a)** density, **b)** richness and **c)** clonality comparisons between thoracic and non-thoracic tumors. T-cell **d)** density, **e)** richness and **f)** clonality comparison between lymph node and non-lymph node tumors.

**Supplementary Figure 4**

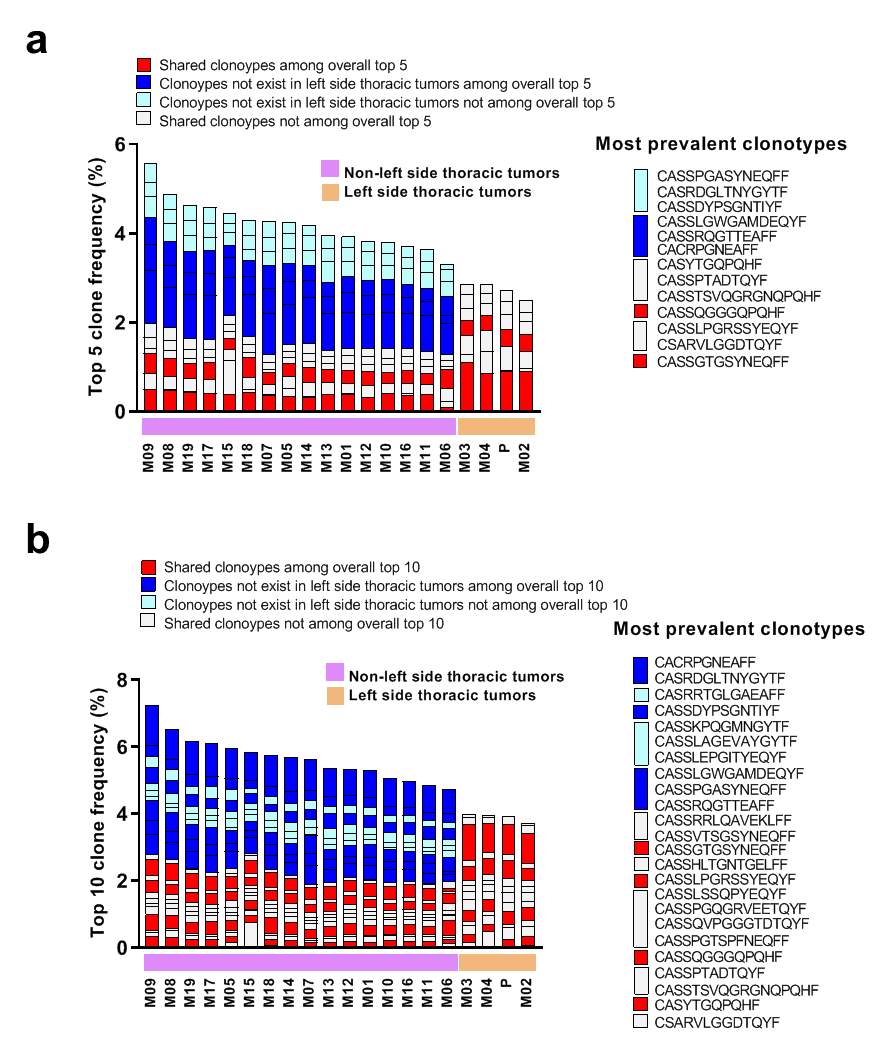

**Supplementary Figure 4. Distinct antigenic profiles are associated with left-side thoracic tumors. a)** Top 5 and **b)** top 10 T cell clonotype distribution landscapes among the 20 tumors.

**Supplementary Figure 5**

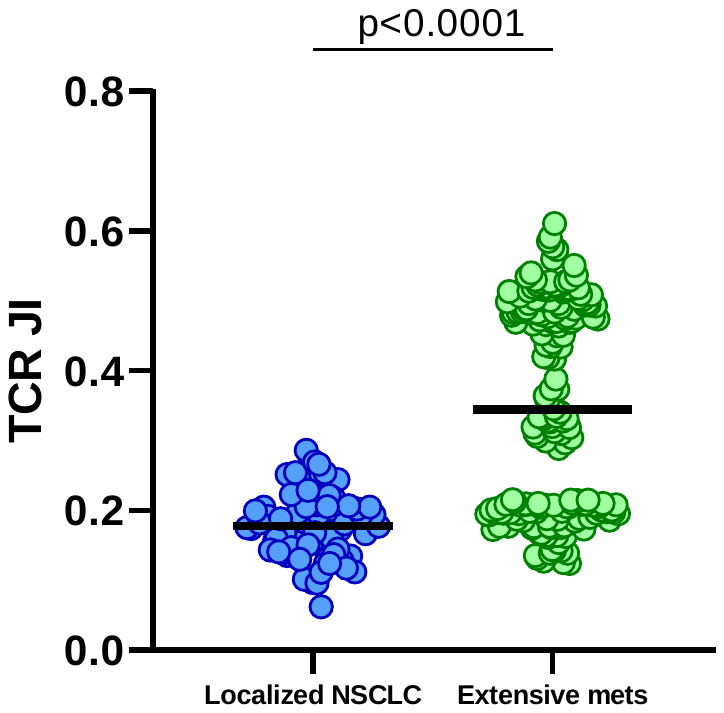

**Supplementary Figure 5. A higher TCR Jaccard index (JI) in our patient compared to a previously published multi-region localized non-small cell lung cancer (NSCLC)**

**Supplementary Figure 6**

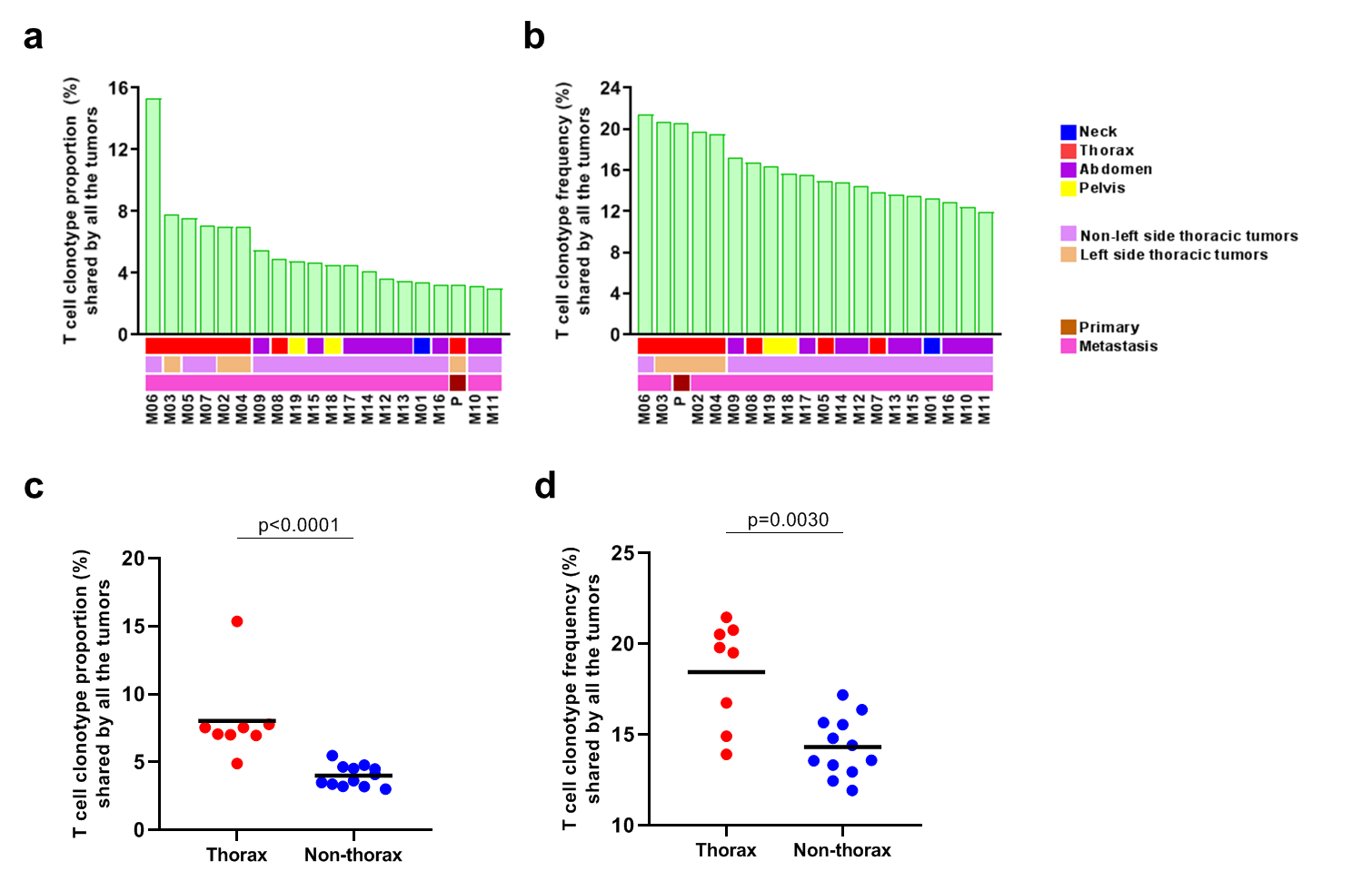

**Supplementary Figure 6. Characterization of shared T cell clonotype proportions and percentages by all 20 tumors.** Shared T-cell clonotype **a)** proportion and **b)** frequency of all 20 tumors. Comparison of shared T-cell clonotype **c)** proportion and **d)** percentage by all the tumors in thoracic and non-thoracic tumor.

**Supplementary Figure 7**

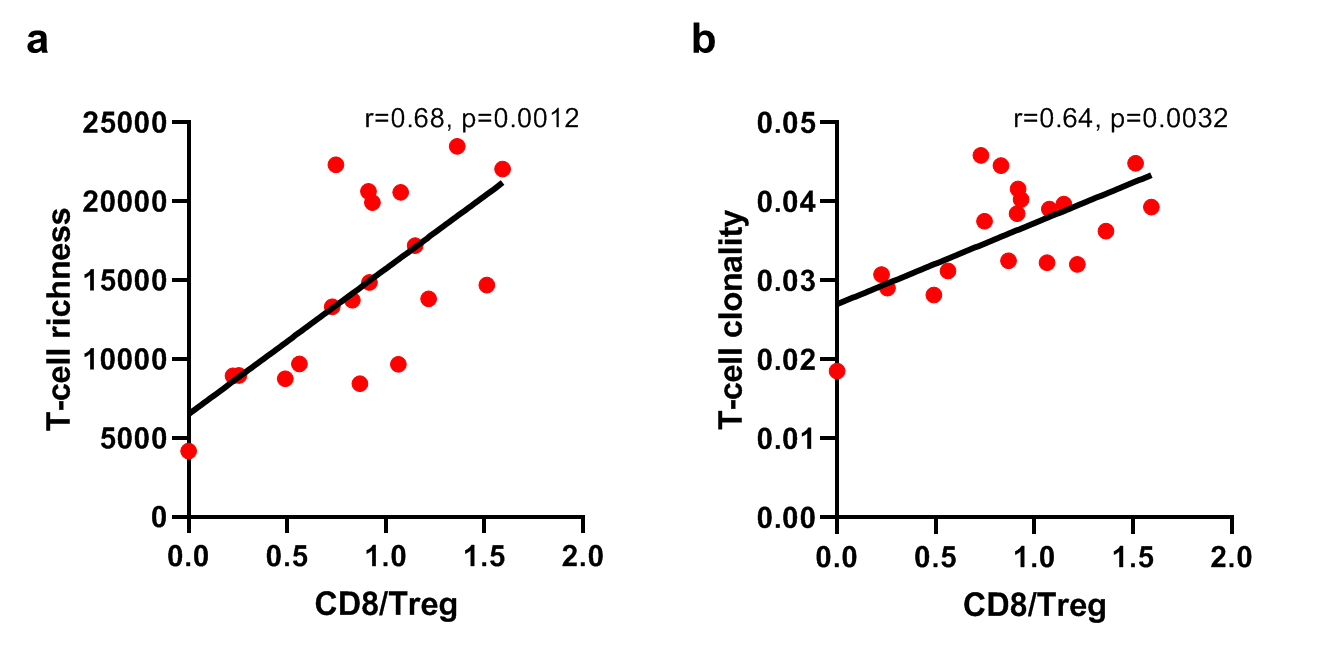

**Supplementary Figure 7. Positive correlations of CD8 to Treg ratio with TCR metrics.** Associations of ratio of CD8 to Treg with T-cell **a)** richness and **b)** clonality.

| **Supplementary table S1. Sample information** | | |  |
| --- | --- | --- | --- |
| Sample ID | Location | System | Described organ |
| M01 | Neck | Endocrine | Thyroid gland |
| P | Thorax | Respiratory | Left lung |
| M02 | Thorax | Respiratory | Left pleural cavity |
| M03 | Thorax | Lymphatic | Left hilar lymph node |
| M04 | Thorax | Respiratory | Left parietal pleura |
| M05 | Thorax | Cardiovascular | Heart |
| M06 | Thorax | Respiratory | Right lung |
| M07 | Thorax | Respiratory | Right pleural cavity |
| M08 | Thorax | Lymphatic | 12^th^ thoracic vertebrate |
| M09 | Abdomen | Digestive | GI tract |
| M10 | Abdomen | Digestive | Liver |
| M11 | Abdomen | Lymphatic | Hepatobiliary lymph node |
| M12 | Abdomen | Lymphatic | Omental lymph node |
| M13 | Abdomen | Lymphatic | Omental lymph node |
| M14 | Abdomen | Lymphatic | Retroperitoneal lymph node |
| M15 | Abdomen | Endocrine | Left adrenal gland |
| M16 | Abdomen | GU | Right kidney |
| M17 | Abdomen | GU | Right kidney |
| M18 | Pelvis | GU | Left ovary |
| M19 | Pelvis | GU | Right ovary |

**Supplementary Data. TCR rearrangements**
